## Supplementary Figures 1 - 11 for "Placental transcription profiling in 6-23 weeks’ gestation reveals differential transcript usage in early development"

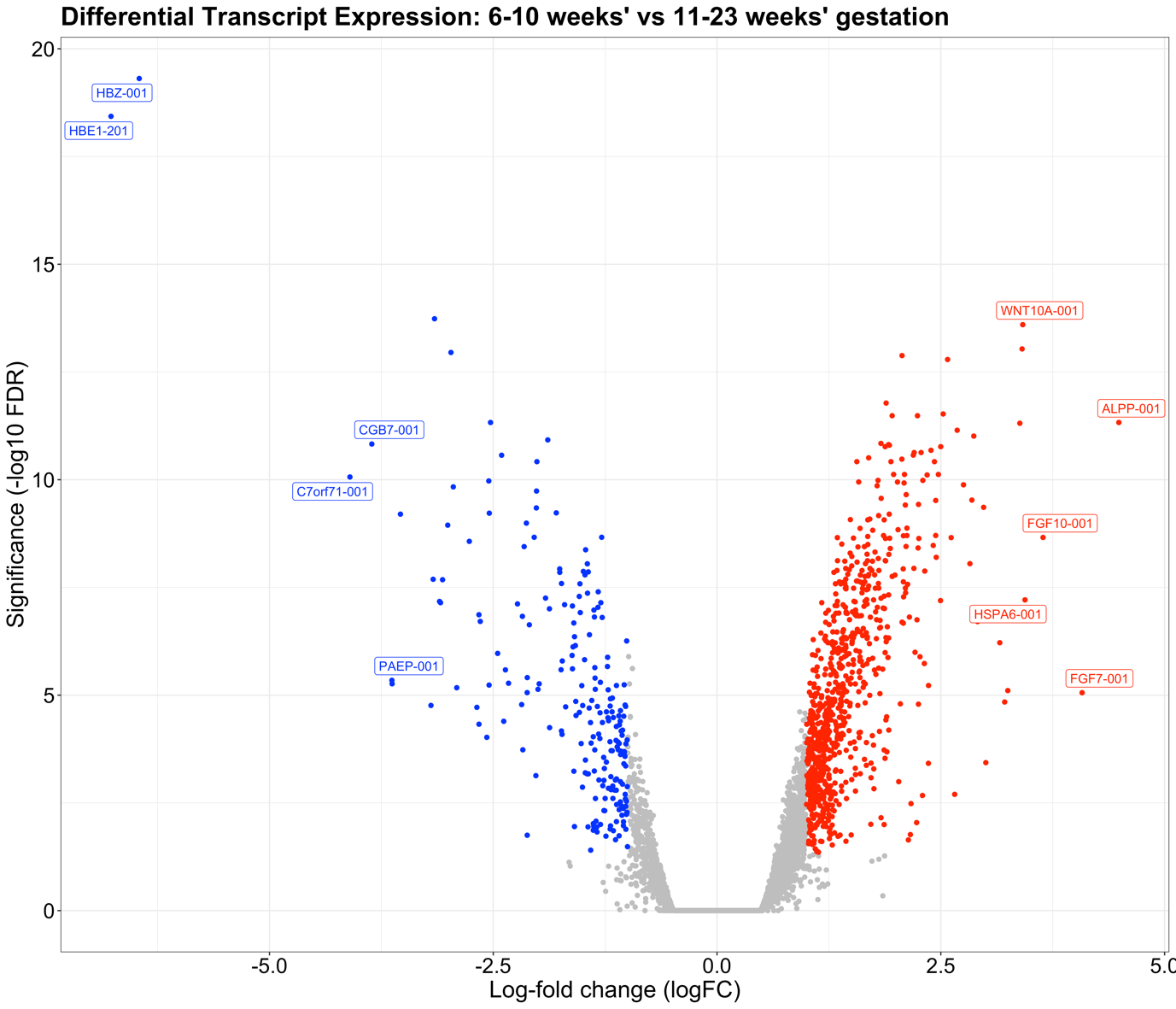

**Figure S1**: Placental villus differential transcript expression compared between 6-10 weeks’ and 11-23 weeks’ gestation. A positive logFC (red) indicates the transcript has a higher expression at 11-23 weeks’ and negative logFC (blue) indicates reduced expression at 11-23 weeks’ compared to 6-10 weeks’ gestation.

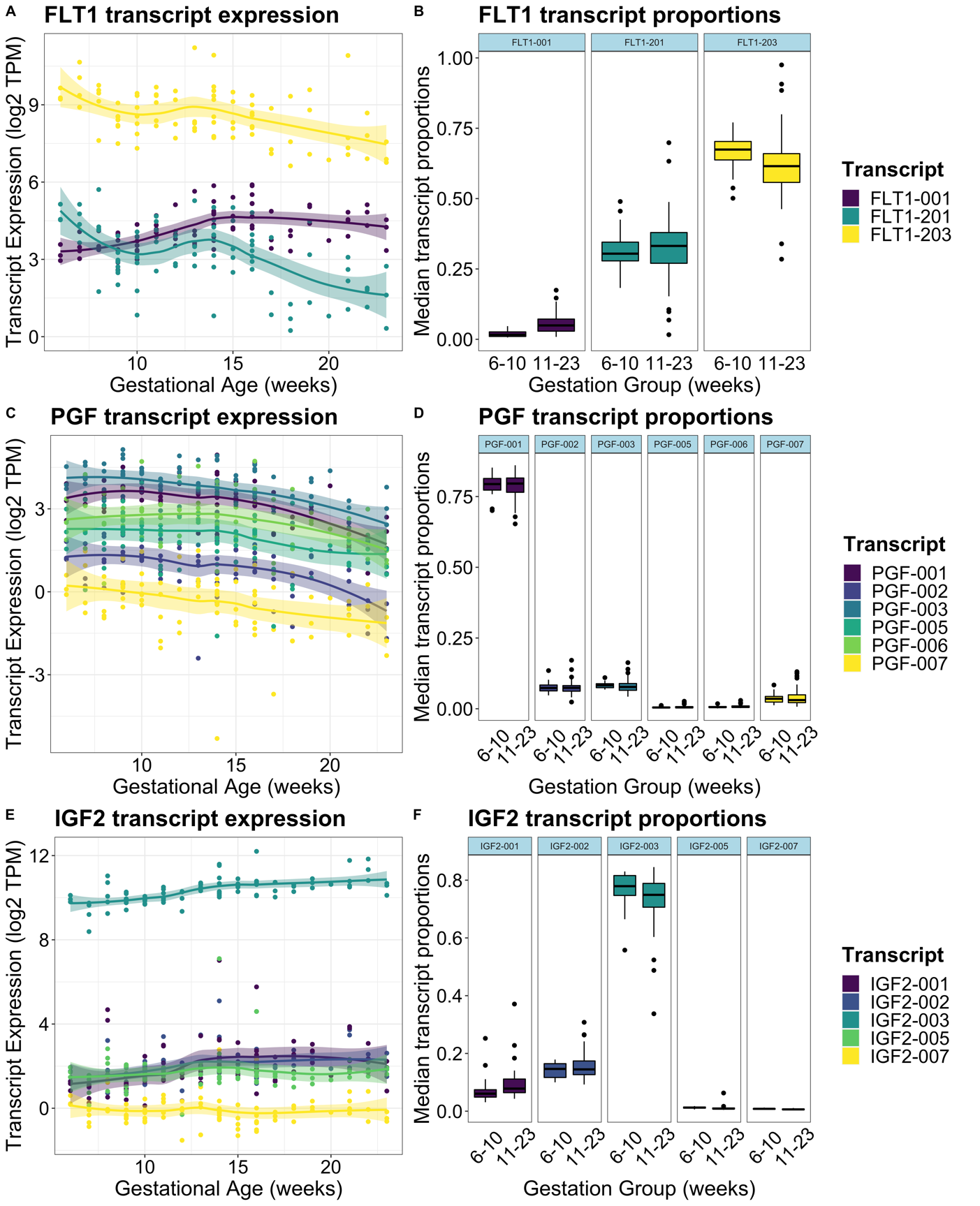

**Figure S2**: Transcript expression in TPM from 6-23 weeks’ gestation (A, C, E) and median transcript proportions between 6-10 weeks’ vs 11-23 weeks’ gestation (B, D, F) of *FLT1*, *PGF*, and *IGF2*. Proportion plots are faceted by individual transcript isoforms.

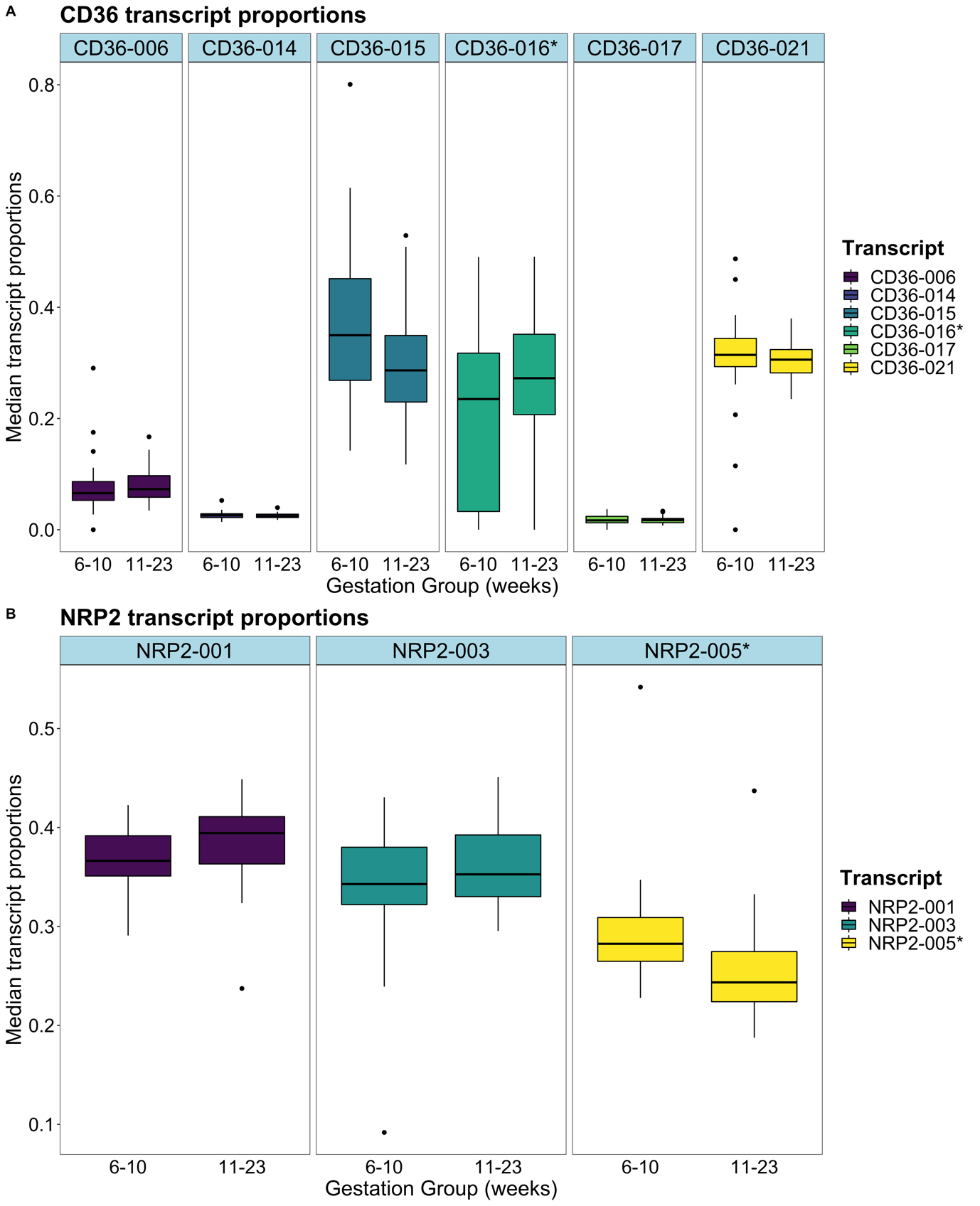

**Figure S3**: Median transcript proportions of *CD36* (A), and *NRP2* (B) between 6-10 weeks’ and 11-23 weeks’ gestation. Both figures are faceted by the individual isoforms present within the genes. Significance is denoted by the asterisk in the strip label and in the figure legend.

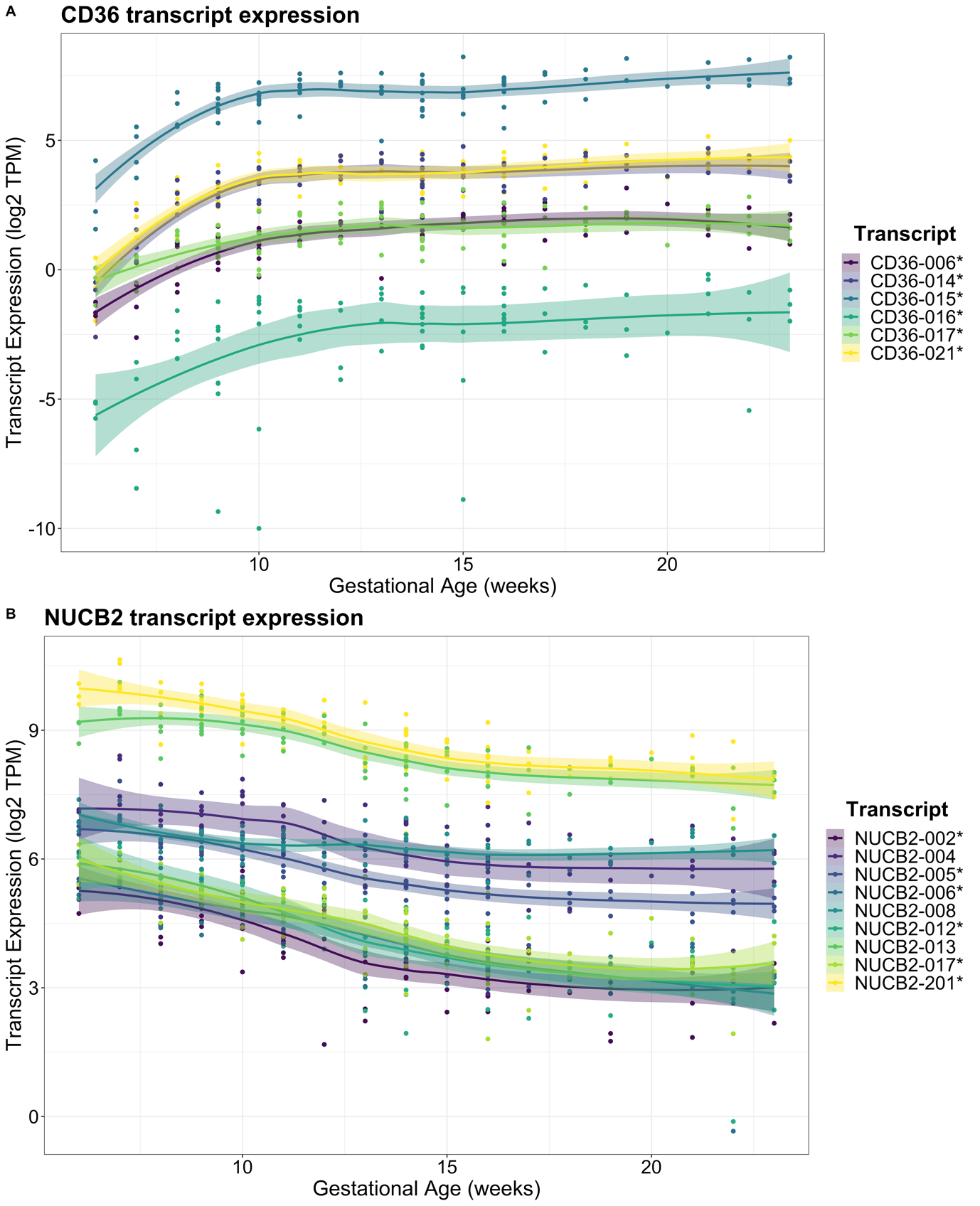

**Figure S4:** Placental villus transcript expression profiles of *CD36* (A) and *NUCB2* (B) from 6-23 weeks’ gestation in log2 transcripts-per-million (TPM) abundances. These genes had the highest numbers of differentially expressed transcripts in placenta across early to mid gestation. Significance is denoted by the asterisk in the figure legend.

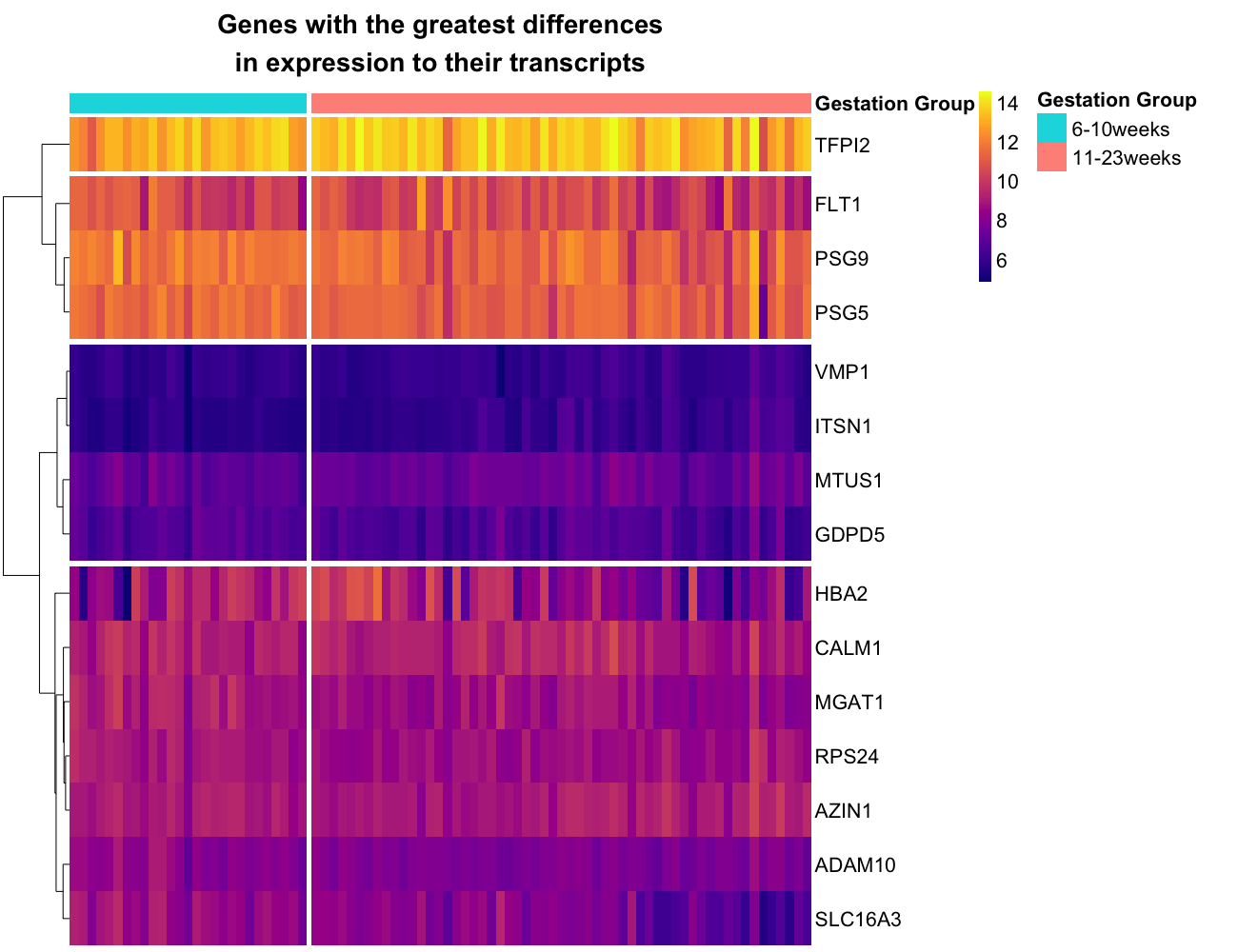

**Figure S5:** Placental villus gene-level expression in log2 counts-per-million (CPM) of the top 15 transcripts shown in figure 3. Rows (genes) are ordered in the same configuration of Figure 3 of the main text. A vertical gap in the heatmap separates the counts into the two sample groups at 6-10 weeks’ and 11-23 weeks’ gestation.

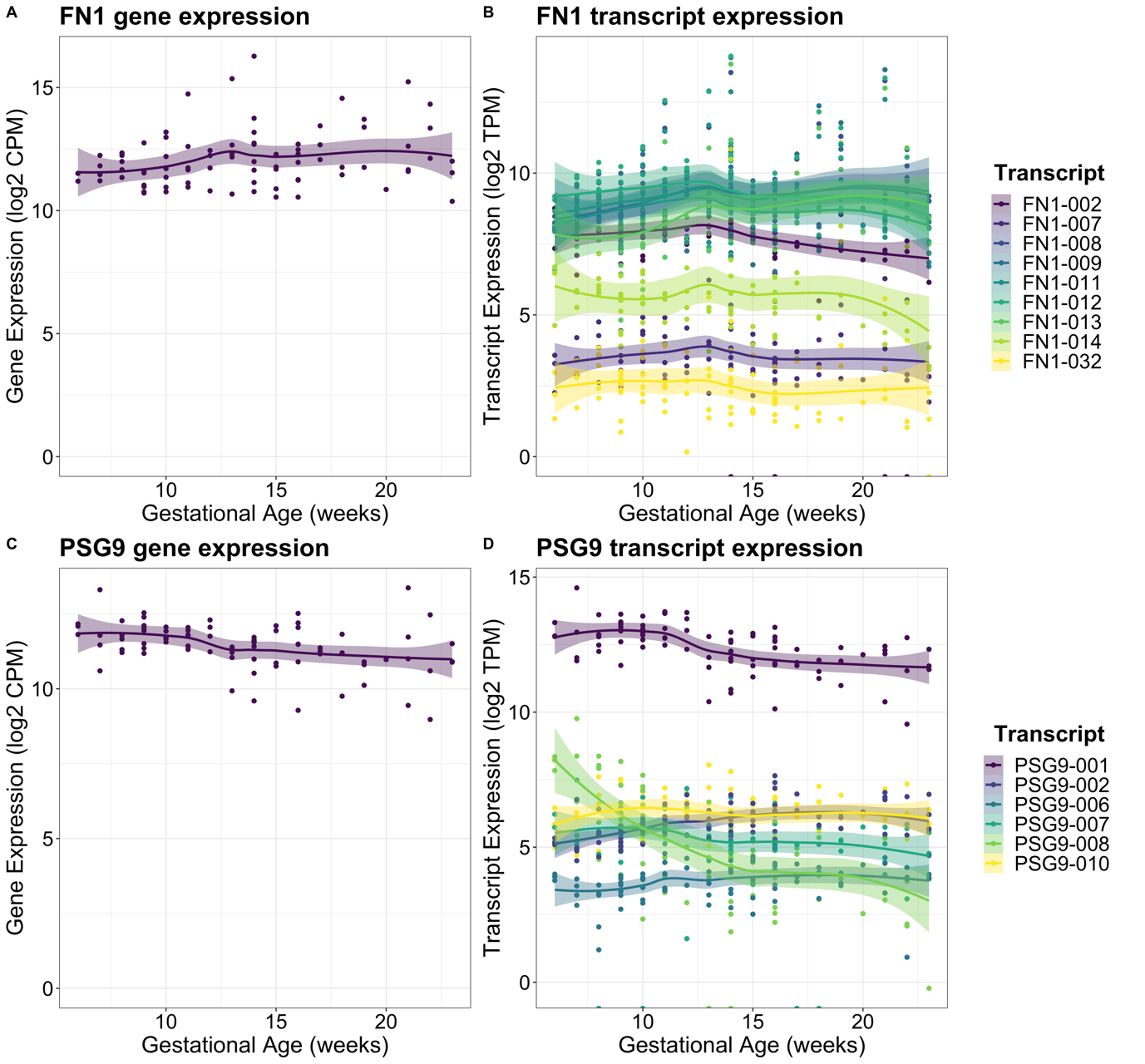

**Figure S6**: Placental villus transcript expression of *PSG9* (B) and *FN1* (D) in log2 TPM abundances, scaled by transcript length, compared to log2 CPM gene expression of *PSG9* (A) and *FN1* (C) across 6-23 weeks’ gestation. Both gene and transcript expression patterns are coloured by transcript.

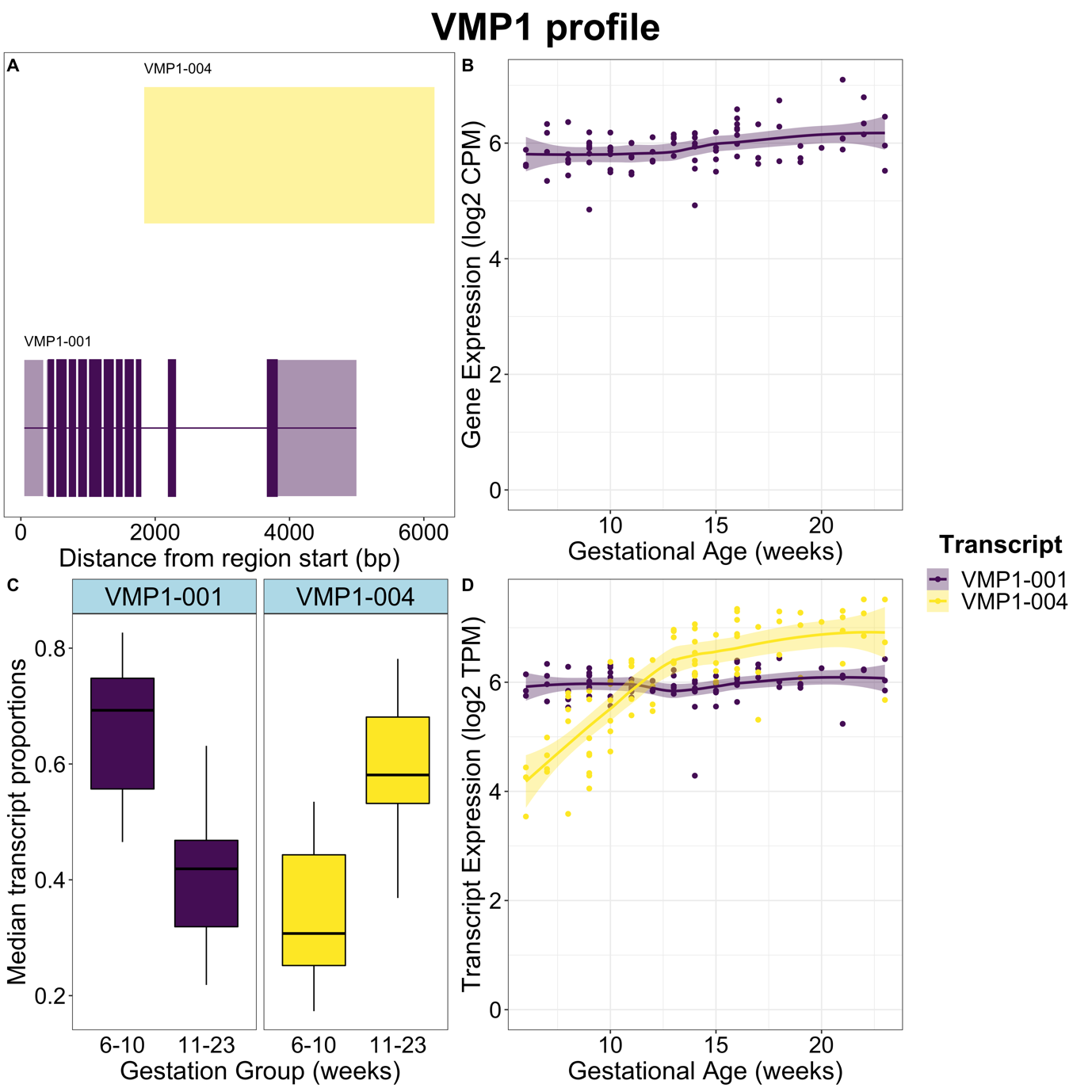

**Figure S7**: Placental villus *VMP1* gene expression, transcript expression, and transcript usage between 6-10 weeks’ vs 11-23 weeks’ gestation, with structures for each transcript shown. A) Transcript structures expressed from the *VMP1* gene. B) Gene expression of *VMP1* in log2 CPM. C) Median transcript proportions of total gene expression in 6-10 weeks’ and 11-23 weeks’ gestation. D) Expression of individual transcripts of *VMP1* in log2 TPM across 6-23 weeks’ gestation.

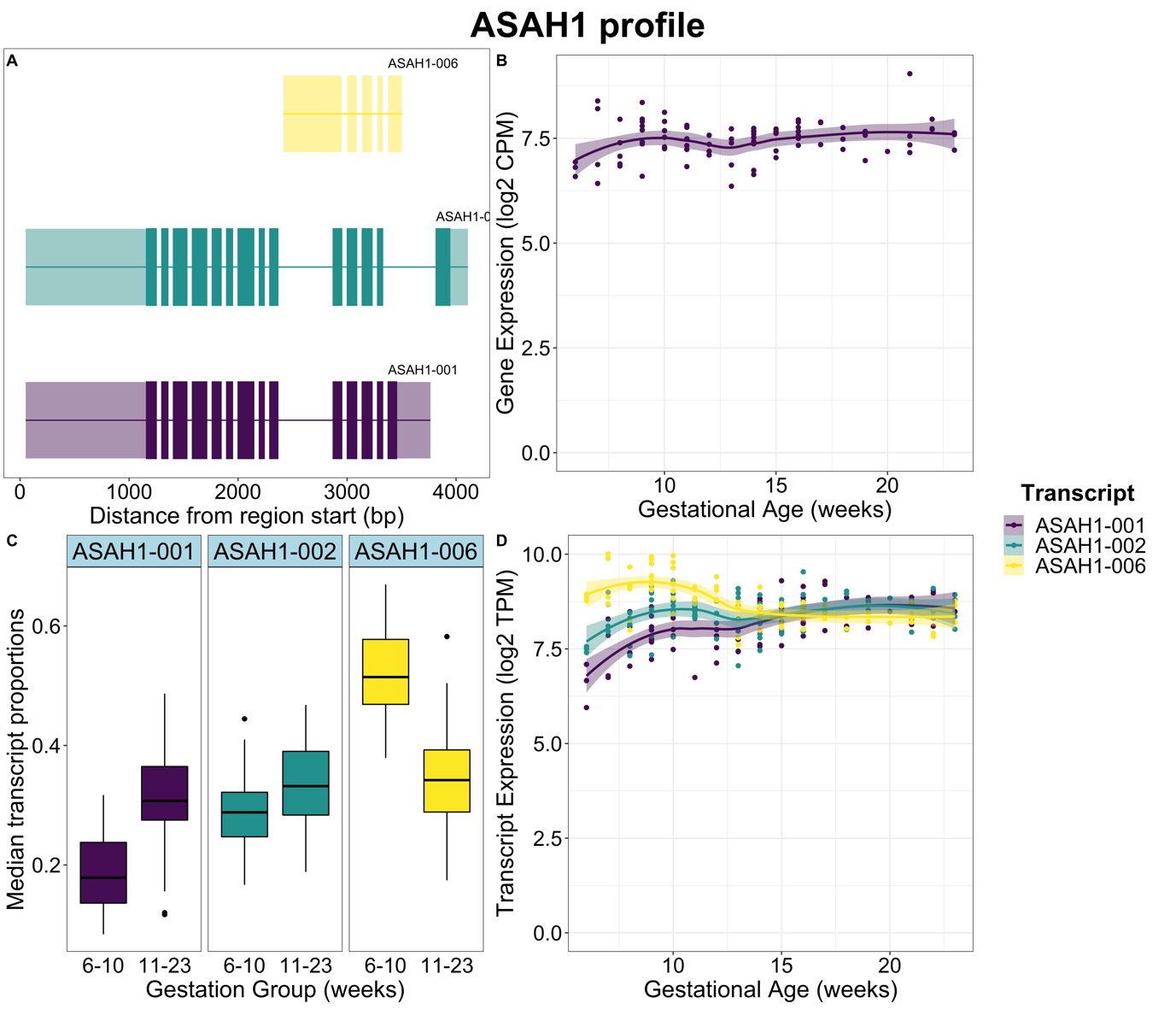

**Figure S8** *ASAH1* gene expression, transcript expression, and transcript usage between 6-10 weeks’ vs 11-23 weeks’ gestation, with structures for each transcript shown. A) Transcript structures expressed from the *ASAH1* gene. B) Gene expression of *ASAH1* in log2 CPM. C) Median transcript proportions of total gene expression in 6-10 weeks and 11-23 weeks’ gestation. D) Expression of individual transcripts of *ASAH1* in log2 TPM.

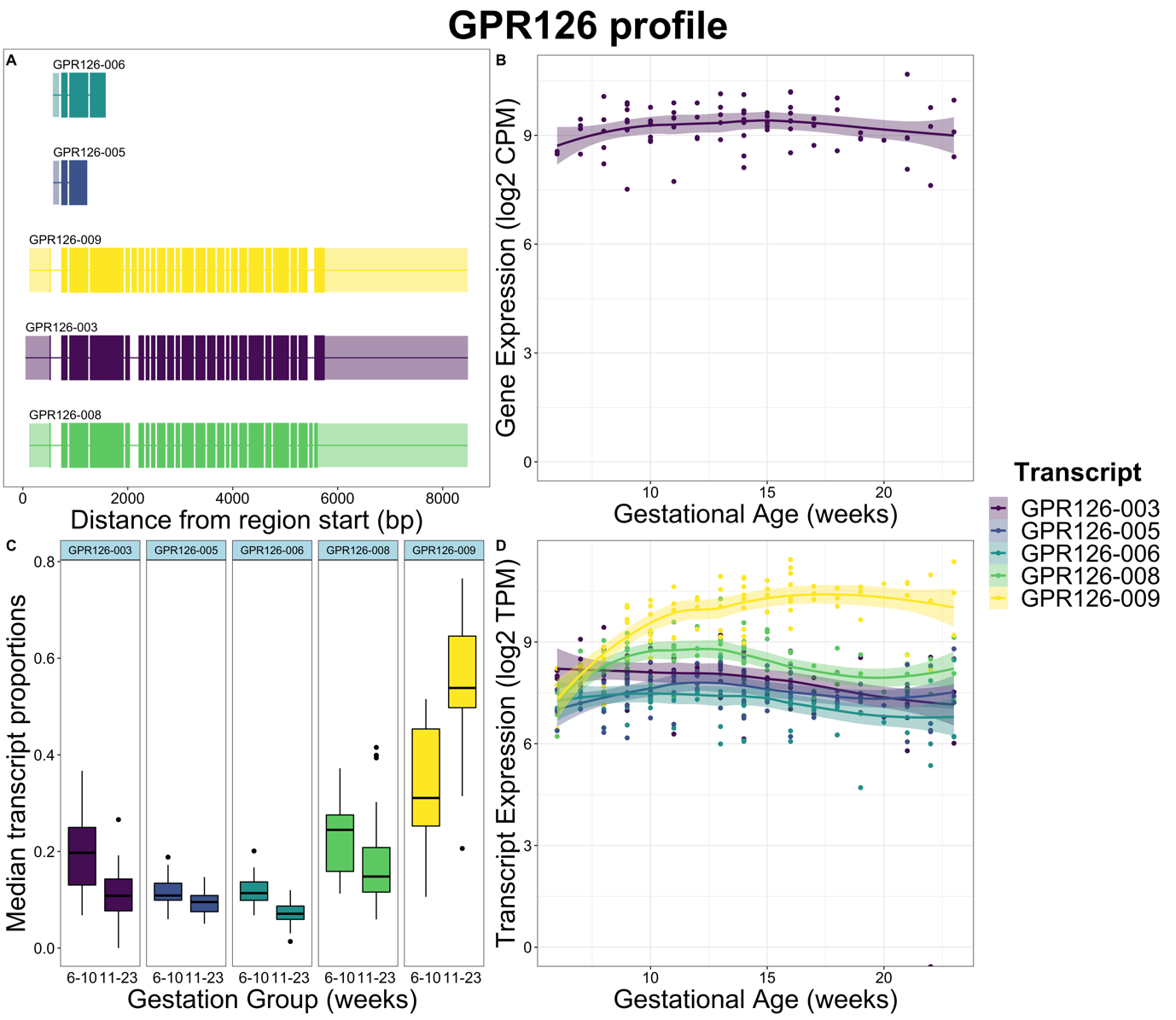

**Figure S9**: *GPR126* (*ADGRG6*) gene expression, transcript expression, and transcript usage between 6-10 weeks’ vs 11-23 weeks’ gestation, with structures for each transcript shown. A) Transcript structures expressed from the *GPR126* gene. B) Gene expression of *GPR126* in log2 CPM. C) Median transcript proportions of total gene expression in 6-10 weeks and 11-23 weeks’ gestation. D) Expression of individual transcripts of *GPR126* in log2 TPM.

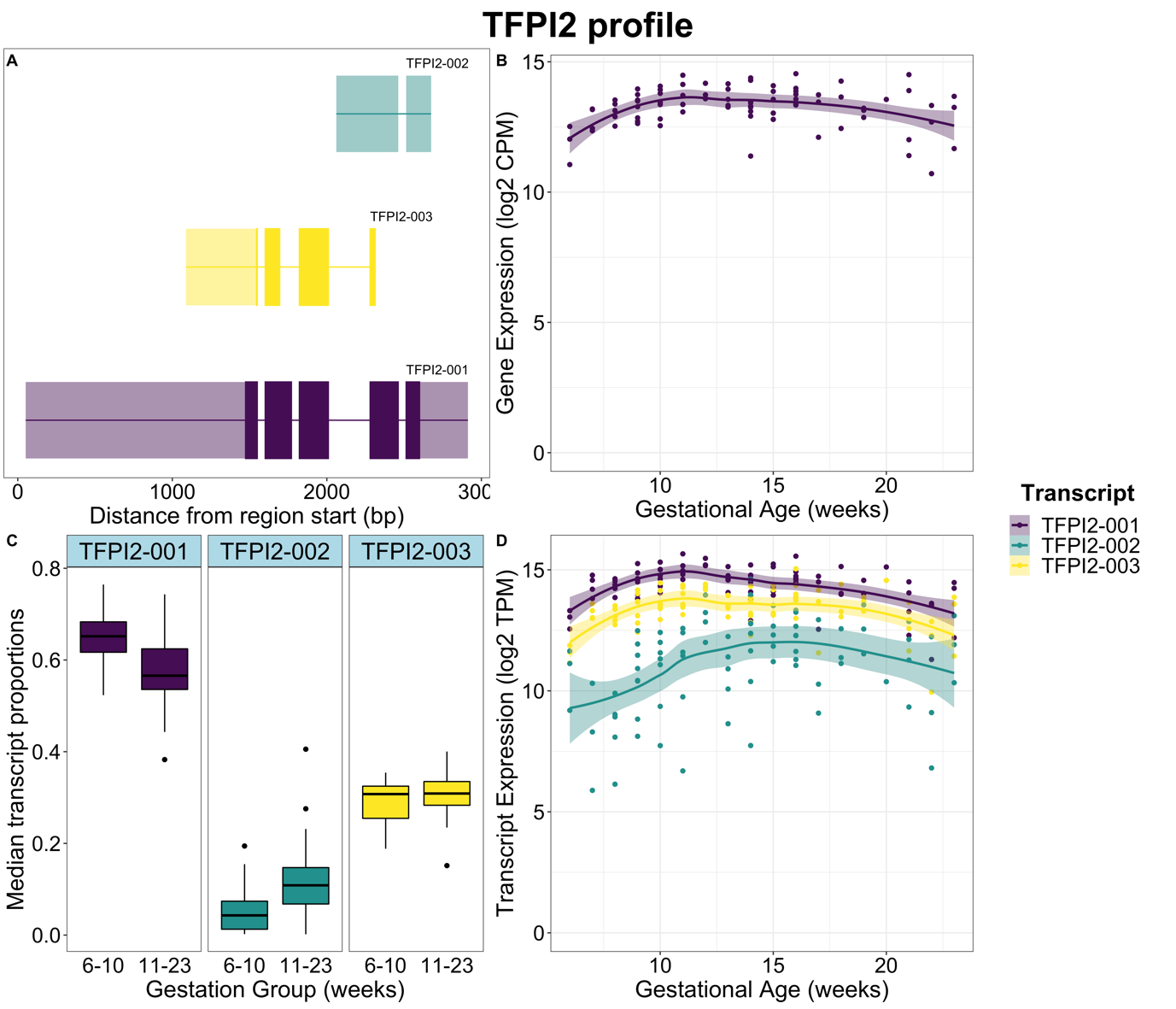

**Figure S10**: *TFPI2* gene expression, transcript expression, and transcript usage between 6-10 weeks’ vs 11-23 weeks’ gestation, with structures for each transcript shown. A) Transcript structures expressed from the *TFPI2* gene. B) Gene expression of *TFPI2* in log2 CPM. C) Median transcript proportions of total gene expression in 6-10 weeks and 11-23 weeks’ gestation. D) Expression of individual transcripts of *TFPI2* in log2 TPM.

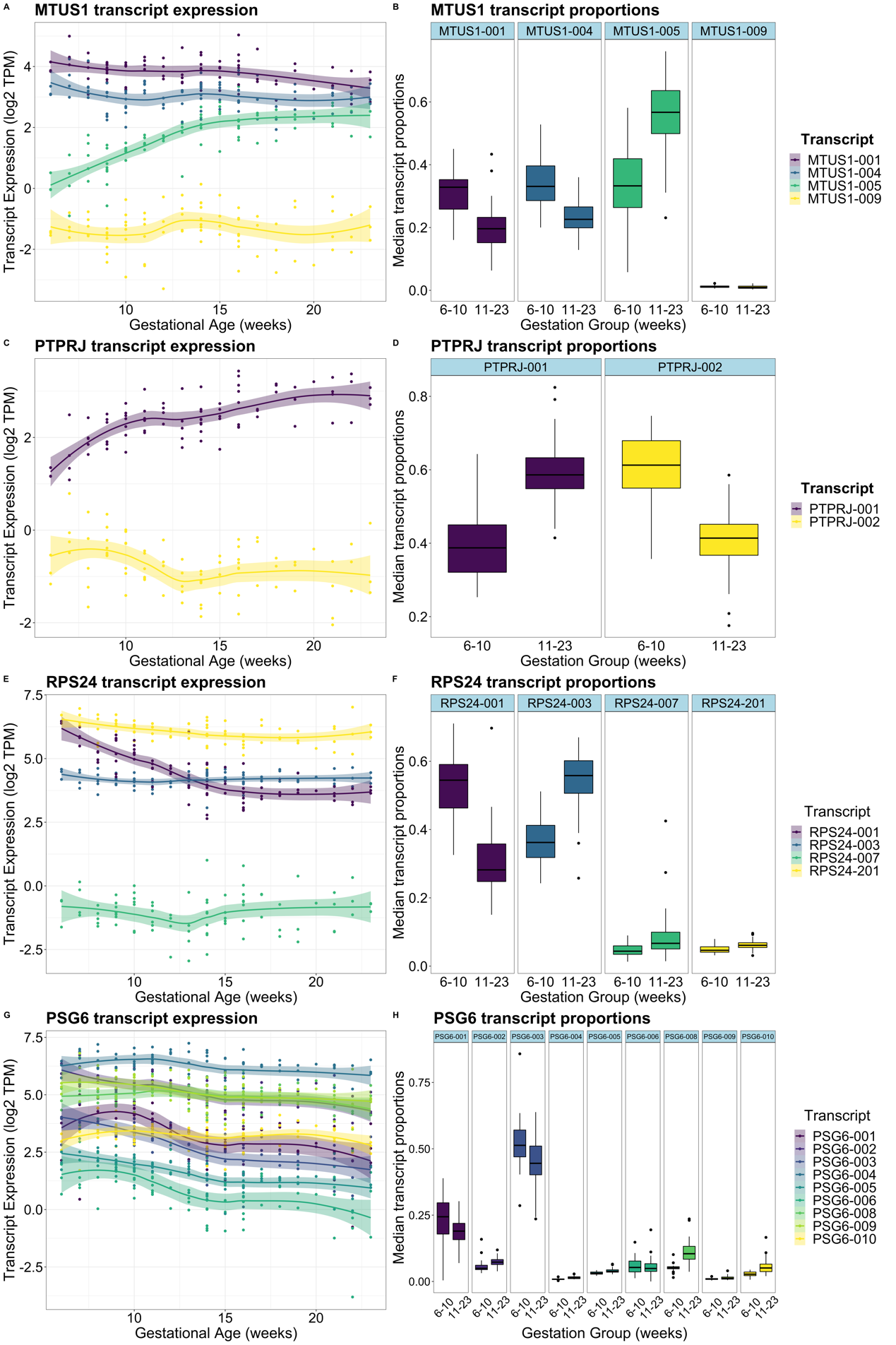

**Figure S11**: Placental villus transcript expression in log2 TPM (A, C, E, G) and median transcript proportions (B, D, F, H) shown for the highest genes in DTU. Median proportions are compared between the two gestational timepoints 6-10 weeks and 11-23 weeks’ gestation, while transcript expression is shown across gestation (6-23 weeks).
